# Cohort Profile Update: Survey of Health, Ageing and Retirement in Europe – Biomarker data for age-related health conditions

**DOI:** 10.64898/2026.01.28.26344911

**Authors:** Martina Börsch-Supan, Axel Börsch-Supan, Karen Andersen-Ranberg, Nis Borbye-Lorenzen, Jake Cofferen, Yacila I. Deza-Lougovski, Rebecca Groh, Solveig Holmgaard, Hannah M. Horton, Elizabeth Kerschner, Thu Minh Kha, Alan J. Potter, Anna Rieckmann, Daniel Schmidutz, Kristin Skogstrand, Aijing Sun, Luzia M. Weiss, Mark H. Wener

## Abstract

SHARE, the “Survey of Health, Ageing and Retirement in Europe”, is the largest population-based panel survey among people aged 50+ in 28 European countries and Israel. It investigates health, economic and social circumstances over the life-course to shed light on the challenges of population ageing. From 2004 until 2023, more than 615,000 in-depth interviews with 160,000 respondents have been conducted in nine survey waves.

Health is crucial to understand ageing. Gold-standard measures are based on blood. SHARE therefore collected dried blood spot (DBS) samples in 12 countries during Wave 6 in 2015. Approximately 27,200 respondents consented (67%).

DBS samples were analysed for overall 21 blood biomarkers in three distinct sets (conventional, cytokine and neurodegenerative biomarkers) between 2017 and 2025. The collection of blood-based health data expands SHARE’s socioeconomic focus with epidemiological insights. SHARE has sparked many collaborations since 2004, and we expect the new biomarker data to inspire further collaborative biomarker projects and data sharing.

## The original cohort

The research infrastructure SHARE (Survey of Health, Ageing and Retirement in Europe) is the largest pan-European social science panel survey to study the health, social, economic, and environmental living conditions over the life-course of citizens in Europe and Israel. It provides internationally comparable longitudinal micro data both for scientists and policy makers. Mission and methodology have been described^1^. From 2004 until 2023, more than 615,000 in-depth interviews with 160,000 respondents aged 50 years or older from 28 European countries and Israel have been conducted in nine survey waves. SHARE has global impact since it not only covers all EU member countries in a strictly harmonised way but additionally is embedded in a network of sister studies all over the world, from the Americas to Eastern Asia. The US Health and Retirement Study (HRS) was the role model for this network of ageing surveys.

## What is the reason for the new data collection?

Health in SHARE has been measured by self-reports which may be unreliable due to lack of knowledge, memory problems by the respondents or other reasons. SHARE has also added functional health measures, such as grip strength and walking speed. However, to assess the prevalence of illnesses that are common in older age, such as diabetes, cardiovascular diseases (CVD) or dementia, additional objective measures are needed. This is especially important in a cross-national and longitudinal survey like SHARE since self-reports and national differences in the survey environment may hamper the intertemporal, international and intercultural comparability. Gold standard objective health data relies on biomarkers measured in blood samples. Hence, SHARE decided to collect dried blood spot (DBS) samples in eleven European countries (Belgium, Switzerland, Germany, Denmark, Estonia, Spain, France, Greece, Italy, Sweden, Slovenia) and Israel during Wave 6 in 2015. Blood collection was strictly harmonised across countries while observing national and EU ethical and legal requirements. DBS samples were analysed for overall 21 blood biomarkers in three distinct sets between 2017 and 2025. The collection of blood-based health data is a major step to expand SHARE’s socioeconomic focus with epidemiological insights.

## What will be the new area of research?

Measuring health is fundamental to understand ageing both on the individual and the population level. It is central for new research in public health as blood marker data complements the health information self-reported by the respondents and will shed light on population health, in particular the prevalence of illnesses common at older ages. It will enhance epidemiological research since individual biomarker values can signal the onset of a disease in its early stage when still unknown to the respondent. It will also open new venues for social and economic research because the new health data will help to overcome biases in international comparisons, such as differential item functioning of self-reported health^2^, thereby more precisely elucidating associations between socioeconomic status and health of individuals as well as the effects of health care systems on population health.

## Who is in the cohort?

Eligibility for DBS collection was restricted to panel households, i.e. all respondents in Wave 6 who had previously participated in any SHARE wave and their partners (the latter independent of previous participation in SHARE), a total of 40,049 respondents. Of these, those who completed a DBS donation consent form were invited to participate. No blood was collected from respondents who were not able to consent themselves. The mean consent rate in the twelve countries was 67.2%, with Belgium, Switzerland, Denmark, Estonia, Sweden, and Slovenia reaching a consent rate over 80%. Across the twelve SHARE countries, 26,358 DBS samples were of a volume sufficiently large for laboratory analysis^3^. To account for potential selection bias in the DBS subsample, we computed DBS-specific weights (Supplement S1 with Figure S1).

## What has been measured?

We measured three sets of biomarkers in the blood material (Table 1). Set 1 includes conventional blood biomarkers, set 2 cytokines and apolipoproteins, and set 3 biomarkers of neurodegeneration.

**Table 1.**
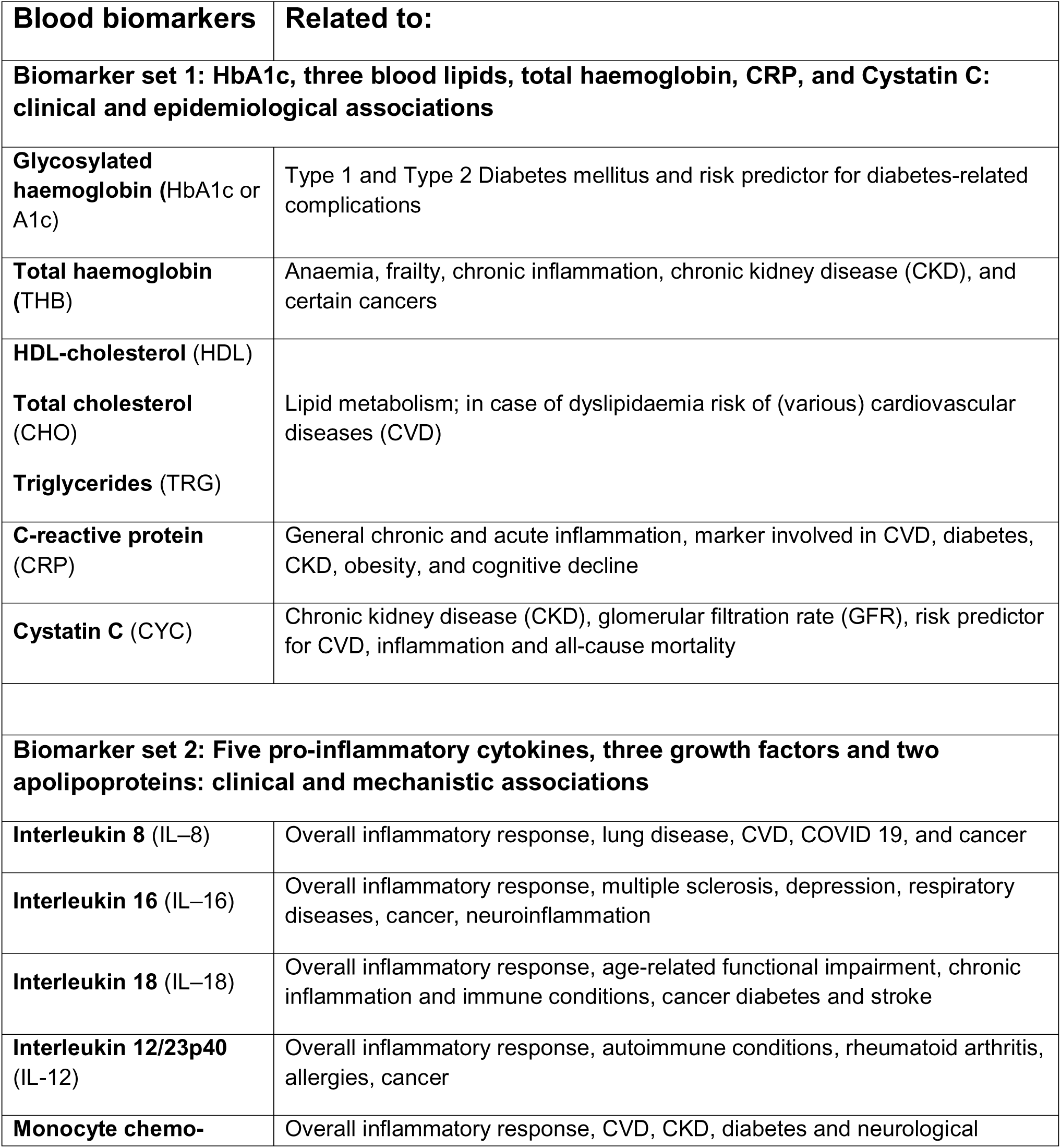
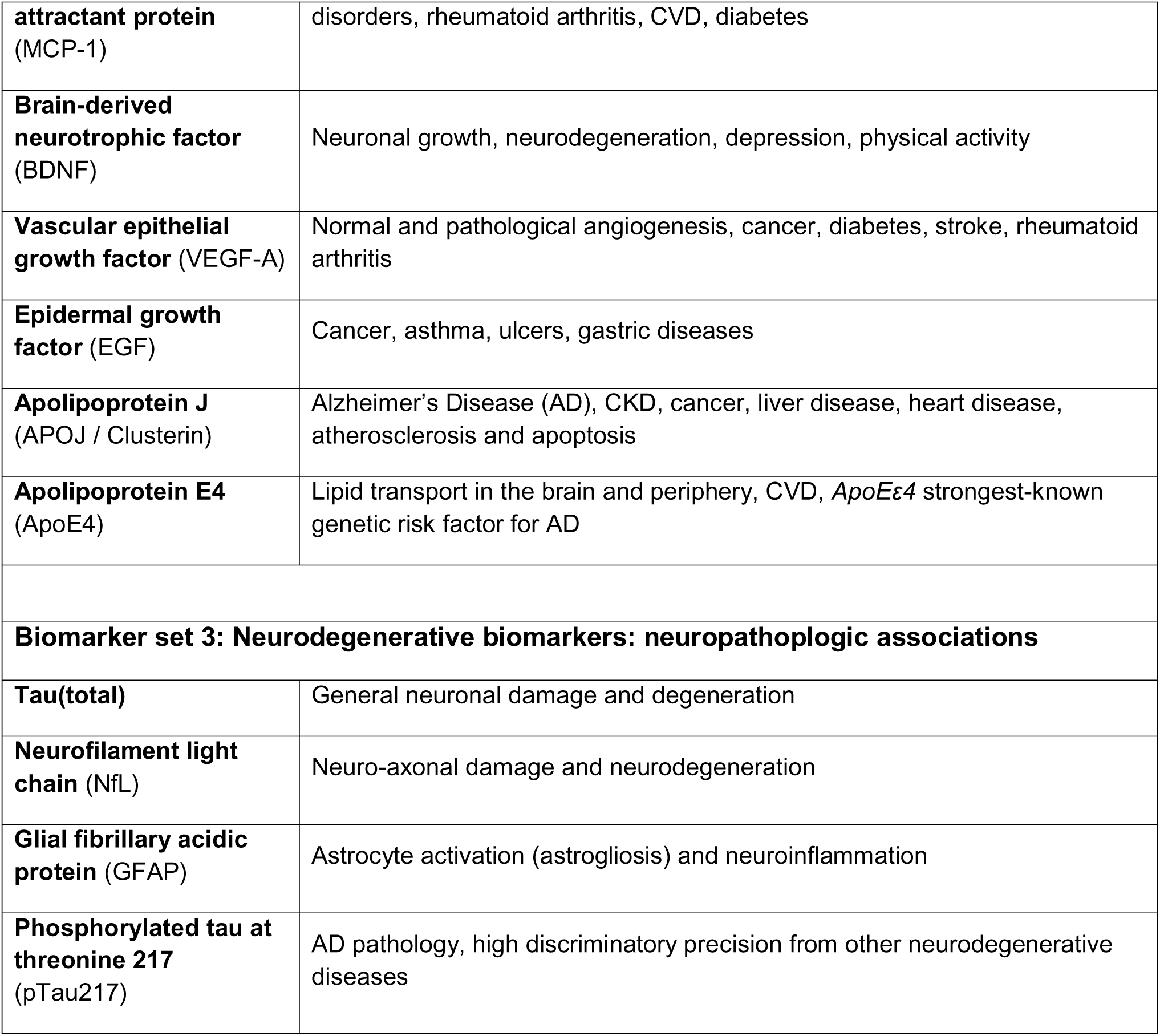
Biomarkers and their main involvement in diseases and age-related conditions.

The measurement followed three steps: first, the collection of dried blood in the respondent’s home and transport to the SHARE biobank, second, the chemical analyses in two different laboratories and third, a statistical adjustment for survey conditions. Details of these three steps are provided in Börsch-Supan et al.^4^.

Step 1: Approval by the ethical committees and obtaining consent is described in Schmidutz and Weiss^5^. During the interview, non-fasting capillary finger-prick blood was collected on DBS cards by trained interviewers and sent as quickly as possible by regular (air)mail to the SHARE Biobank (Department of Public Health, University of Southern Denmark, Odense, Denmark), for freezer storage at −23°C until analyses and thereafter. The monitoring of data collection and a description of the division of the SHARE DBS into subsamples (punches) can be found in Börsch-Supan et al.^4^.

Step 2: Not all DBS samples could be assayed immediately due to funding limitations and lab closures during the COVID pandemic. In 2018, one-third of set 1 was assayed at the Department of Laboratory Medicine and Pathology, University of Washington (UW), Seattle, WA, USA. The analyses of the remaining two-thirds of biomarker set 1 followed in 2020/21 at UW. The assay protocols including SHARE-specific modifications of biomarker set 1 are described in Supplement S2 and^4,6,7^. All samples of sets 2 and 3 were assayed at the Statens Serum Institut (SSI) in Copenhagen, Denmark in 2018 and 2025, respectively (Supplements S3 and S4, Supplementary Tables S3.1 and S4.1).

Step 3: During sample collection in a non-clinical setting, DBS are inevitably exposed to varying fieldwork conditions, such as differences in outside temperature, humidity and shipping time^8,9,10^. Sample quality may also be affected by abbreviated DBS drying times, missing humidity protection during shipment or collection of smaller than optimal blood volumes^7^. Degradation of the biomarkers due to extended storage as well as changes of the analysing instruments in the interim between the earlier and later analyses of set 1 required adjustment of the biomarker levels^6,7,11,12^. The subset of DBS samples analysed for neurodegenerative markers (set 3) in 2025 had an especially long storage time since collection in 2015. Finally, as capillary blood eluted from DBS is different from venous blood, the biomarker values needed an adjustment for specimen-type to permit results obtained from analyses of DBS to be evaluated using metrics established for venous blood. This adjustment was based on laboratory simulation studies^6^ and was applied to the biomarkers in set 1 for which well-established standard values are available (Supplement S5, with Table S5.1 and Figure S5.1).

Biomarker set 1 (Conventional blood biomarkers): Set 1 included blood biomarkers related to diseases common in older age and routinely measured at doctor visits, such as blood levels of glycosylated haemoglobin (HbA1c) for diabetes; total haemoglobin (THB) for, e.g. anaemia, frailty; blood lipids triglycerides (TRG), total and HDL-cholesterol (CHO and HDL) for cardiovascular diseases; CRP for inflammation status; and cystatin C (CYC) for kidney function. The DBS subsamples (s. *Step 1*) were extracted and blood levels of these markers were determined at UW in 2017/18 and 2020/21. Assays were conducted according to published techniques, but adapted for the SHARE-specific analyses of multiple biomarkers using the eluent from only a subsample of one or two punches (Supplement S2). Reference values from venous blood samples are well established for each of these markers. The blood values of the biomarkers of set 1 were released to the SHARE users in 2024^4^.

Biomarker set 2 (Cytokines, growth factors and apolipoproteins): Set 2 has been designed to better understand the complex relationship between biology, health behaviour and socioeconomic status in the ageing process. Among the underlying factors that cause ageing-related diseases, including cognitive decline and neurodegeneration, are inflammatory pathways, neoplastic growth or genetic risk factors. Cytokines (IL-8, IL-16, IL-18, IL12/23, MCP-1) are small blood-based proteins prominently involved in the inflammatory processes, atherosclerosis, CVD, obesity, diabetes; growth factors (BDNF, VEGF-A, EGF) stimulate benign cell growth and differentiation as well as malignant growth of neoplasms; the apolipoproteins ApoE4 and clusterin are involved in (mild) cognitive decline and dementia/Alzheimer’s Disease (AD). Moreover, the *ApoE*_ε_*4* gene variant is the strongest known genetic risk factor for AD. These markers are not routinely analysed in a blood count, but in research they offer the opportunity to investigate different inflammation processes or fine-tune the search for risk factors of CVD and cognitive decline. Using multiplex immunoassay analysis, multiple markers can be analysed from very small amounts of blood. Borbye-Lorenzen & Börsch-Supan^13^ describe the selection of the ten markers and the methods of their analysis. The blood level values of these markers were established at SSI in 2018. Laboratory processing and validation are described in Supplement S3 with Table S3.1. Differently from the biomarkers of set 1, there are no standard values available.

Biomarker set 3 (Neurodegenerative biomarkers): Set 3 complements the psychometric measurement of cognition that has been in SHARE since its first wave. It has been motivated by the rising number of individuals with mild to severe cognitive impairment which imposes an increasing burden not only on the individuals and their families but also on healthcare systems and economies. Based on research in recent years^e.g.,14,15,16,17^, we selected the neurodegenerative biomarkers GFAP, NfL, Tau(total), and pTau217 for further analysis of the remaining DBS from respondents in Germany, Denmark and Sweden. The laboratory analyses for these biomarkers were performed at SSI in 2025 (see Supplement S4 with Table S4.1).

## What has been found? Key findings and publications

Table 2 presents the main descriptive statistics of the 21 biomarkers. Not all SHARE respondent DBS samples have observations for all markers. For samples with limited blood spot volume, priority of analyses was given to the analysis of biomarkers of high-risk diseases and health conditions common at older ages, such as cardiovascular diseases, diabetes and chronic kidney disease (CKD). Biomarkers of set 3 were analysed from remaining DBS material in Denmark, Germany and Sweden.

**Table 2.** Descriptive statistics of biomarkers.

| Marker | Unit | Obs. | Mean | Median | Std. Dev | Min | Max |
| --- | --- | --- | --- | --- | --- | --- | --- |
| **Set 1: Seven routine blood biomarkers** |  |  |  |  |  |  |  |
| Glycosylated haemoglobin (HbA1c) | % | 19,788 | 6.0 | 6.0 | 0.6 | 3.9 | 16.5 |
| Total haemoglobin (THB) | g/dL | 14,902 | 14.8 | 14.7 | 1.3 | 9.8 | 22.0 |
| HDL-cholesterol (HDL) | mg/dL | 14,902 | 68.5 | 67.9 | 9.2 | 37.9 | 121.9 |
| Total cholesterol (CHO) | mg/dL | 23,061 | 223.0 | 223.1 | 24.9 | 118.3 | 337.7 |
| Triglycerides (TRG) | mg/dL | 22,989 | 223.0 | 196.7 | 127.6 | 9.8 | 3,386 |
| C-reactive protein (CRP) | mg/L | 22,728 | 3.5 | 2.1 | 6.6 | 0.1 | 139.4 |
| Cystatin C (CYC) | mg/L | 23,061 | 1.0 | 1.0 | 0.1 | 0.5 | 2.5 |
| **Set 2: Pro-inflammatory cytokines and growth factors** |  |  |  |  |  |  |  |
| Interleukin 8 (IL-8) | pg/mL | 15,512 | 3.0 | 2.5 | 7.2 | 0.4 | 515.4 |
| Interleukin 16 (IL-16) | pg/mL | 15,435 | 327.2 | 315.1 | 111.0 | 34.8 | 2,651 |
| Interleukin 18 (IL-18) | pg/mL | 15,515 | 110.3 | 103.9 | 41.6 | 7.1 | 726.6 |
| Interleukin 12/23p40 (IL-12) | pg/mL | 15,384 | 12.2 | 9.6 | 8.9 | 0.5 | 139.5 |
| Monocyte chemo-attractant protein (MCP-1) | pg/mL | 15,515 | 46.2 | 44.5 | 14.9 | 7.7 | 565.9 |
| Brain-derived neurotrophic factor (BDNF) | pg/mL | 15,214 | 40.9 | 38.2 | 18.0 | 12.2 | 286.7 |
| Vascular epithelial growth factor (VEGF-A) | pg/mL | 15,514 | 14.1 | 13.3 | 5.00 | 3.2 | 83.3 |
| Epidermal growth factor (EGF) | pg/mL | 15,231 | 3.2 | 2.9 | 1.7 | 0.1 | 24.5 |
| **Set 2 cont'd: Apolipoproteins** |  |  |  |  |  |  |  |
| Apolipoprotein J (Clusterin) | ng/mL | 15,514 | 1,778 | 1,686 | 526.3 | 32.6 | 8,266 |
| Apolipoprotein E4 (ApoE4) | ng/mL | 13,108 | 62.8 | 7.2 | 105.7 | 3.7 | 1,192 |
| **Set 3: Neurodegenerative biomarkers** |  |  |  |  |  |  |  |
| Tau(total) Tau(total) | pg/mL | 3,103 | 10.8 | 10.1 | 4.4 | 2.25 | 55.0 |
| Neurofilament light chain (NfL) | pg/mL | 2,488 | 3.7 | 2.7 | 3.9 | 0.15 | 61.3 |
| Glial fibrillary acidic protein (GFAB) | pg/mL | 3,054 | 1.3 | 1.1 | 1.1 | 0.085 | 33.3 |
| Phosphorylated tau at threonine 217 (pTau217) | pg/mL | 2,921 | 2.2 | 1.8 | 2.1 | 0.094 | 50.3 |

We found substantial country differences and often a north-south gradient within Europe (Figure 1 and Supplement 6 with Table S6). This holds after adjusting for field conditions, among them the temperature differences between north and south. THB, HDL and CHO are highest in Sweden and lower in all Mediterranean countries (Italy, Spain, Greece, and lowest in Israel). CYC follows that pattern with the exception of Italy. CRP is highest in Spain, Germany and Denmark, lowest in Greece. TRG is highest in France and Switzerland and lowest in Denmark and Estonia. HbA1c is highest in Greece and lowest in Denmark with a remarkable north-south gradient across the twelve SHARE countries. The ApoE4 protein also exhibits a strong north-south gradient, a finding in accordance with previous studies on the distribution of the *ApoE*_ε_*4* gene variant^18^.

**Figure 1.**
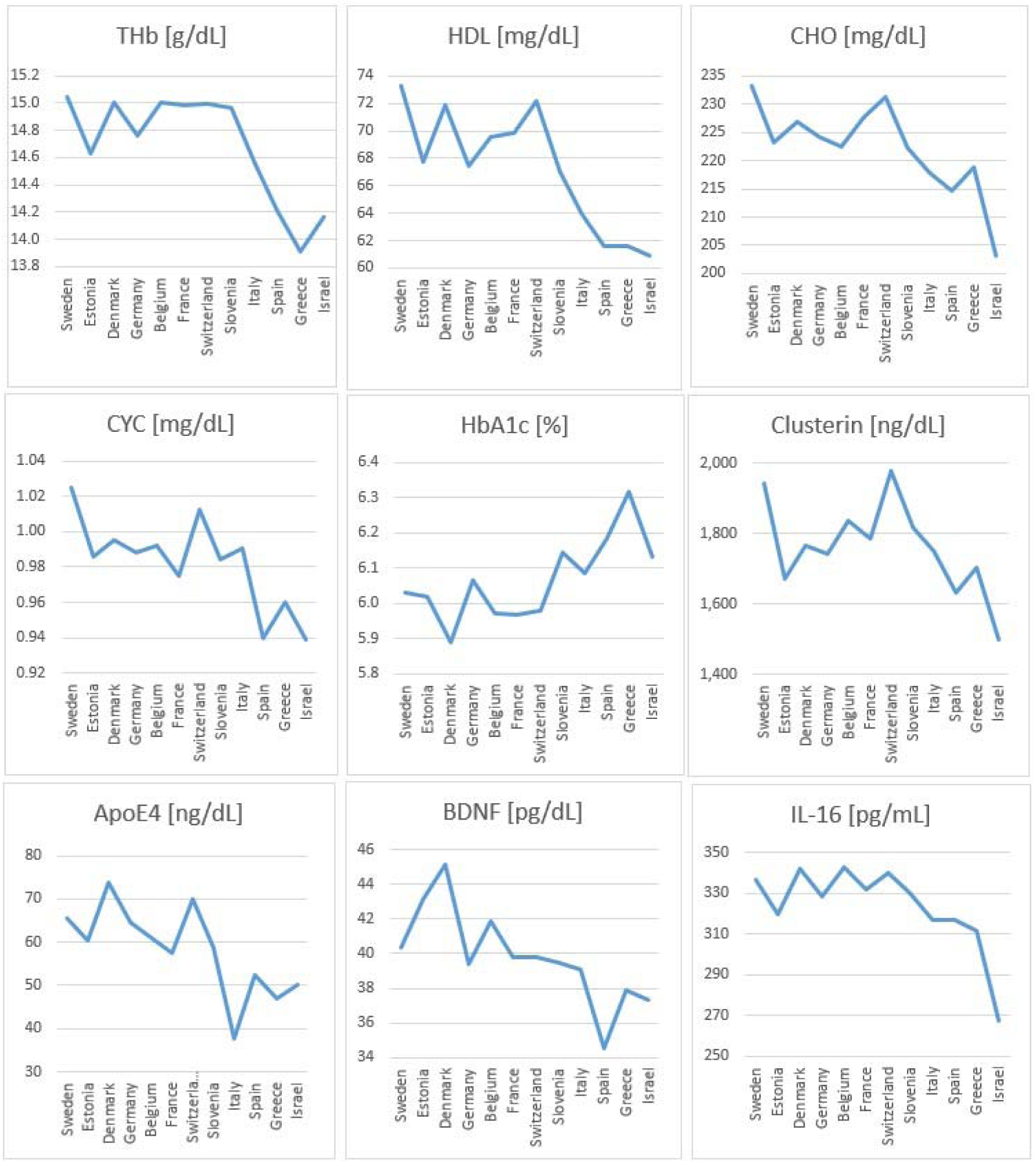
Variation of selected biomarkers by country

Asking SHARE respondents during the interview for diseases diagnosed by a doctor, we were able to monitor self-reporting or detect conditions like diabetes or CKD unknown to the respondent. We found that a large share of respondents was unaware that they had one of these common age-related illnesses. Blood sugar levels measured from the SHARE DBS collection revealed that 7.2% of all respondents have HbA1c values larger than 6.5% (the percentage defining diabetes) but did not report that a doctor ever diagnosed the disease. This represents 34% of all diabetes cases^19^ and is disconcerting as long-term healthcare costs for diabetes are very high and can be prevented with early detection and lifestyle changes.

Similarly, we found substantial unawareness of CKD; 11% of the respondents did not report a diagnosis of CKD although it was indicated by their cystatin C blood level. One might expect that individuals with hypertension, diabetes or a large number of comorbidities would more frequently report a diagnosis of CKD. This was not the case. Even among respondents with four or more comorbidities, the likelihood of reporting a CKD diagnosis was only about 27%^20^.

Borbye-Lorenzen et al.^21^ employed a genetic analysis to match the ApoE4 protein level determined from our DBS samples with carrying at least one *ApoE*_ε_*4* allele. This is the case for 24% of our participants^21,18^. Furthermore, Deza-Lougovski et al.^22^ showed that *ApoE*_ε_*4* carriers in the SHARE sample are significantly more likely to have cognitive impairment. For the neurodegenerative markers analysed from blood collected in 2015, Börsch-Supan et al.^23^ found evidence for associations between pTau217, GFAP and NfL and cognitive performance in 2022 in SHARE Wave 9 seven years later. pTau217 has the strongest significance and, with few exceptions, also the strongest effect size.

## What are the main strengths and weaknesses?

The main strength of the data is its objectivity which overcomes the self-reporting biases often inherent in subjective health data and self-reported doctor diagnoses. Collection and analyses of DBS were harmonised, enabling international comparability of blood biomarker data. Validation experiments were performed to correct for the impact of field conditions (present in non-clinical environment) on the collected blood biomarker levels. We could demonstrate that ApoE4 protein is easy to collect in a population survey and can be used in lieu of the genetic data^21^. Moreover, it is an important biomarker when researching cognitive decline in larger populations. In this context and despite long storage time, the remaining blood material from three countries allowed the assessment of neurodegenerative blood markers. The results encourage larger-scale collection and analysis of these markers from DBS to use their predictive power of cognitive decline to implement lifestyle changes as early as possible for possible postponement of dementia onset.

The main weakness is the limitation to only twelve of the 28 countries in SHARE, and to only three countries for the neurodegenerative markers. Missing baseline levels and standard blood values for the biomarkers of sets 2 and 3 allow only in-sample comparisons. Another weakness is the sensitivity to survey conditions such as temperature, humidity, drying and shipping time, and spot size. This requires elaborate validation methods (biomarker set 1) and post-collection statistical adjustments (sets 2 and 3). The long storage times may have had adverse effects on the biomarker levels especially for sets 2 and 3.

## Can I get hold of the data? Where can I find out more?

The biomarker data is available as a special module of the SHARE data [https://share-eric.eu/data/data-set-details/share-dbs]. Access to the SHARE data is provided free of charge to all scientists globally. Details on how to register as a SHARE user are available at www.share-eric.eu. The central SHARE Coordination Team provides user support under: [https://share-eric.eu/data/faqs-support]. Scientific support for the biomarker data will be provided by the Munich Research Institute for the Economics of Aging and SHARE Analyses (MEA) [https://www.mea-share.eu/dbs/]. Contact Martina Börsch-Supan.

## Ethics approval

The data collection in the Survey of Health, Ageing and Retirement in Europe (SHARE) procedures are subject to continuous ethics review. SHARE-ERIC’s activities related to human subjects’ research are guided by international research ethics principles such as the Respect Code of Practice for Socioeconomic Research (professional and ethical guidelines for the conduct of socioeconomic research) and the ‘Declaration of Helsinki^24^. The collection of blood samples during Wave 6 in 2015 was approved by the ethics committees of all participating countries and the Ethics Council of the Max Planck Society. Obtaining informed written consent from the respondent was obligatory in all participating countries and this consent could be revoked at any time even after blood collection. The necessary procedures are documented in several publications and reports^4,5,25,26^.

## Author contributions

MBS oversaw the biomarker project. ABS coordinated SHARE and was responsible for funding. MBS and ABS had primary responsibility for writing the manuscript. LMW and MBS designed and organised the data collection processes and the validation of biomarker set 2. ABS and LMW designed the validation experiment to adjust for survey conditions for biomarker set 1. DS took care of ethics and data protection issues. KAR coordinated the SHARE biobank. JC, EK, TMK, AJP, and MHW executed the laboratory analyses of marker set 1 at UW in Seattle, WA, USA. NBL and KS organised and executed punching for all markers and laboratory analyses for marker sets 2 and 3 at SSI in Copenhagen. RG developed the algorithm for spot size correction. AS created the DBS specific weights. ABS, AR, YIDL, SH, HMH, and LMW performed the regression analyses for our findings. All authors participated in drafting and critically reviewing the manuscript.

## Acknowledgements

We thank the central SHARE Coordination Team, the country teams and the survey agencies of the SHARE countries that participated in the DBS collection. Our special appreciation goes to both the respondents who donated their blood and the interviewers who conducted the blood collection in the field.

## Supplementary Material

### Supplement S1: Construction of weights

The lack of eligibility and consent, plus the failure to produce blood spots that are sufficiently large to be analysed for at least one biomarker may lead to sample selectivity, i.e., the risk of bias due to systematic differences between eligible and non-eligible respondents, consenters and non-consenters, and analysed and non-analysed DBS samples (Figure S1). To account for this selection, we computed DBS weights, which researchers should use when they analyse the DBS data. The DBS weights are the product of calibrated cross-sectional individual weights from the main SHARE survey Wave 6 (cciw_w6) and a correction factor for the lack of eligibility, consent or analysability.

**Supplementary Figure S1:**
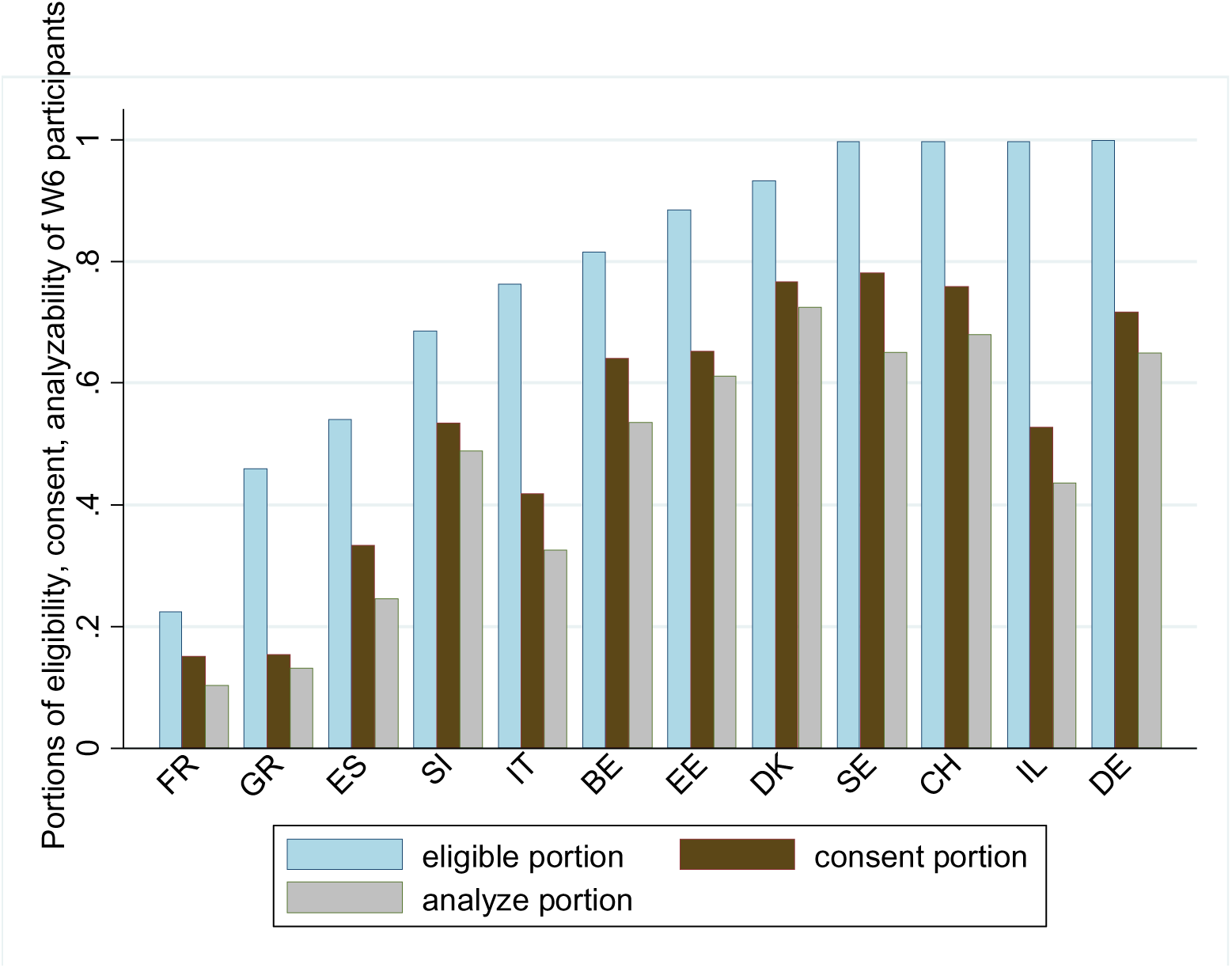
Eligibility, consent and analysability.

For this correction factor, we use an inverse probability weighting method. We estimate a logistic model to predict the probability of having at least one biomarker value among all SHARE W6 participants in these 12 countries who completed interviews. The predictor variables for this model are demographic characteristics (gender, age, education) and country fixed effects, specified as dummy variables with *K* categories. Moreover, we included the interaction between education and gender. The dependent variable *DBS_ij_* indicates that individual *i* in country *j* has at least one biomarker value.

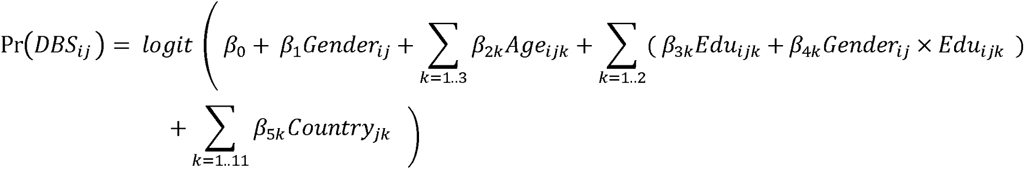

The regression was weighted with the baseline calibrated cross-sectional individual weights *cciw_w6* to match the population distribution of the predictors. Gender and education did not significantly affect DBS participation. Compared with respondents aged 50-59, respondents aged 60-79 are more likely to provide a DBS sample. Compared with Germany, people in Denmark and Switzerland are more willing to provide blood samples and have samples which meet the analysis criteria. All other countries have a lower probability to provide analysable DBS when compared to Germany. The predicted inverse probability from the logit regression was multiplied with the calibrated cross-sectional individual weights *cciw_w6* from SHARE Wave 6 to obtain the DBS weights:

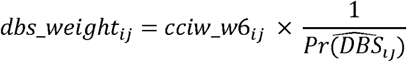

### Supplement S2: Description of SHARE dried blood spot (DBS) assays of biomarker set 1 performed at the, Department of Laboratory Medicine and Pathology, University of Washington, Seattle, USA

#### Methodology

Gold-standard assays were performed on liquid (i.e., serum, plasma, and/or whole blood) specimens to establish the target concentrations of analytes in calibrators, quality control (QC) specimens, and assay validation specimens. Total cholesterol (CHO), triglycerides (TRG), HDL cholesterol (HDL), and C-reactive protein (CRP) concentrations were measured by an AU680 Chemistry Analyzer (Beckman Coulter, Miami, FL, USA).

Total haemoglobin (THB) concentration was measured by a Sysmex XE-5000 hematology analyzer (Sysmex, Mundelein, IL, USA). Glycosylated haemoglobin A1c (HbA1c) percentage was measured by a Variant II and/or D100 ion-exchange high-performance liquid chromatography system (HPLC; Bio-Rad Laboratories, Hercules, CA, USA), and, for confirmation of HbA1c values, by an immunoturbidimetric method run on a UniCell DxC 600 Chemistry Analyzer (Beckman Coulter). Cystatin C (CYC) concentration was measured by a sandwich enzyme-linked immunosorbent assay (ELISA; BioVendor, Candler, NC, USA).

The DBS assays for CHO, TRG and HDL were modified versions of the gold standard enzymatic methods. In brief, a filter paper disc(s) punched from a DBS card was eluted with a buffer solution to elute (re-liquefy) the dried blood. An aliquot of DBS eluate was incubated with an assay-specific enzyme to produce hydrogen peroxide, which then reacted with horseradish peroxidase (HRP) and a fluorogen to produce a fluorescent fluorophore. The fluorescent intensity of the solution, proportional to the concentration of analyte in the specimen, was measured by a spectrophotometer (BioTek, Winooski, VT, USA).

The DBS assays for CRP and CYC were modified versions of well-established sandwich ELISA methods. For those two assays, in brief, an aliquot of DBS eluate was incubated in a reaction plate coated with an antibody specific to the biomarker of interest. The plate was then washed, a detector antibody coupled to HRP was added, excess HRP was washed away, chromogenic substrate was added, and light absorbance proportional to the amount of analyte in the specimen was measured by a spectrophotometer (BioTek). HbA1c was measured in DBS eluates by HPLC and/or by immunoturbidimetry. THB concentration was determined by incubating an aliquot of DBS eluate with 12.5% Triton X-100/0.1% sodium hydroxide (NaOH), and absorbance was then measured by a spectrophotometer (BioTek). Assay-specific details are given below.

#### Calibrators and Quality Control (QC)

Liquid plasma assay calibrators were constructed from high concentration pooled human plasma (UWLMP) serially diluted with 7% bovine serum albumin in phosphate buffered saline (BSA/PBS; 74 Sigma Aldrich, St. Louis, MO, USA) to the desired final analyte concentrations. 50µL aliquots of each calibrator were pipetted onto CF12 filter paper (Whatman, Piscataway, NJ, USA), dried for four hours at room temperature (RT), sealed in Ziploc bags with desiccant packs, and stored at −80°C to create dried plasma assay calibrators. Two DBS QC samples (i.e., sample controls) were constructed to have assay-specific clinically-relevant high and low analyte concentrations by mixing pools of human plasma and washed red blood cells in equal volumes (i.e., 50% haematocrit), pipetted in 75µL aliquots onto CF12 filter paper, dried for four hours at RT, sealed in Ziploc bags with desiccant packs, and stored at −80°C. Prior to spotting, the analyte concentrations of the liquid calibrators and QC samples were determined by analysis using the gold-standard methods described above.

#### Assay Details

To accommodate the limited material available from SHARE DBS, the samples were analysed using modified versions of UWLMP standard DBS assays. HDL, THB, and HbA1c (these jointly extracted markers were designated *A-markers* as an interim identifier during analyses) were measured in shared eluent obtained from two 3.2mm diameter discs punched from a DBS. CHO, TRG, CRP, and CYC (designated *B-markers* as an interim identifier during analyses) were measured in shared eluent obtained from a single 3.2mm disc punched from a DBS. For SHARE DBS with very limited material, HbA1c alone was measured in eluent obtained from a single 3.2mm disc punched from a DBS (Am1punch).

##### A-markers

Two 3.2mm discs were eluted by shaking for one hour at RT in 400µL double-distilled water (ddH2O). A 200µL aliquot of the eluate was transferred to a pierceable microvial followed by addition of 550µL ddH2O to a final volume of 750µL, and HbA1c then measured by HPLC using the proprietary software provided with the instrument (Bio-Rad). To 200µL of the remaining initial eluate, 200µL ddH2O was added to a final volume of 400µL. 40µL of that solution was incubated with 75µL Reagent 1 (Beckman Coulter) for 30 minutes at 37°C. 50µL of Reagent 2 (Beckman Coulter), diluted 1:1 with ddH2O and containing 0.01% 10-acetyl-3,7-dihydroxyphenoxazine (ADHP; Cayman Chemical, Ann Arbor, MI, USA), was then added. After 30 minutes incubation at 37°C, the fluorescence emission of the solution was measured at 590nm using 540nm excitation by a Synergy HT spectrophotometer (BioTek). Gen5 software (BioTek) was used to construct a standard curve by plotting the fluorescence values of the standards against the known HDL concentrations. Using the standard curve, the fluorescence values of the DBS were interpolated to determine HDL concentrations of the samples. THB was measured by incubating 50µL of the above 400µL DBS eluate solution with 100µL 12.5% Triton X-100/0.1% NaOH (Sigma Aldrich) for 15 minutes at RT and reading absorbance at 405nm using a spectrophotometer (BioTek). A standard curve was constructed by plotting the absorbance values of the standards against the known THB concentrations. Using the standard 75 curve, the absorbance values of the DBS were interpolated to determine THB concentrations of the samples. To measure HbA1c immunoturbidimetrically, a single 3.2mm punch was eluted in 300µL Haemolyzing Reagent (Beckman Coulter, Inc.) by shaking at 1000rpm for 1 hr. at RT. An aliquot of 200µL was transferred to a 0.3mL sample cup and loaded onto the DxC 600 (Beckman Coulter) and assayed using the HbA1c-reagent kit (Beckman Coulter A87905) without modification beyond the DBS sample type.

##### B markers

One 3.2mm diameter disc was eluted by shaking for one hour at RT in 400µL PBS/0.1% Triton X 100/0.01% Tween-20. Total cholesterol reagent (Synermed) was prepared following manufacturer directions and 0.01% ADHP was then added. CHO was measured by combining 20µL DBS eluate and 100µL reagent, incubated for 30 minutes at 37°C, and fluorescence read on a spectrophotometer as given above for HDL. Triglycerides reagent (Beckman Coulter) was prepared by mixing the contents of cartridge A and C and bringing the pH to 7.4. Just prior to assaying, 5% v/v cartridge B and 0.01% ADHP was added to the A/C mixture. TRG was measured by combining 40µL sample with 100µL reagent, incubating for 30 minutes at 37°C, and fluorescence then read on a spectrophotometer as given above for HDL. ELISAs were used to measure CRP (BioCheck Inc., South San Francisco, CA, USA) and CYC (BioVendor) by assaying 80µL and 100µL DBS eluate, respectively, with each assay performed following ELISA manufacturer directions. Am1punch (HbA1c only) one 3.2mm disc was eluted by shaking for one hour at RT in 400µL ddH2O. 350µL ddH2O was then added for a final volume of 750µL, and HbA1c measured by HPLC (Bio-Rad).

#### Assay Performance

##### A markers and HbA1c

The HDL within-assay imprecision (CV) was 7.4% and between-assay imprecision was 11.5%. The HDL concentrations of 50 DBS-paired serum and plasma samples assayed on the AU680 correlated with the HDL concentrations of capillary blood DBS and EDTA venous blood DBS (R^2^ = 0.77 and 0.72, respectively). The THB within-assay imprecision (CV) was 5.2% and between-assay imprecision was 7.4%. The THB concentrations of 50 DBS-paired blood samples assayed by Sysmex correlated with the THB concentrations of capillary blood DBS and EDTA venous blood DBS (R^2^ = 0.59 and 0.43, respectively). The HbA1c within-assay imprecision (CV) was 1.3% and between-assay imprecision was 2.4%. The HbA1c values of 50 DBS-paired blood samples assayed on the HPLC correlated with the HbA1c values of capillary blood DBS and EDTA venous blood DBS (R^2^ = 0.94 and 0.96, respectively). The HbA1c values of 36 DBS-paired blood samples correlated with the HbA1c values of capillary DBS and assayed on the DxC 600 (R^2^ = 0.95).

##### B markers

The CHO within-assay imprecision (CV) was 6.2% and between-assay imprecision was 8.9%. The CHO concentrations of 50 DBS-paired serum and plasma samples assayed on the AU680 correlated with the CHO concentrations of capillary blood DBS and EDTA venous blood DBS (R^2^ = 0.73 and 0.73, respectively). The TRG within-assay imprecision (CV) was 5.3% and between-assay imprecision was 13.7%. The TRG concentrations of 49 DBS-paired serum and plasma samples assayed by the AU680 correlated with the TRG concentrations of capillary blood DBS and EDTA venous blood DBS (R^2^ = 0.94 and 0.89, respectively). The CRP within-assay imprecision (CV) was 4.2% and between-assay imprecision was 11.9%. The CRP concentrations of 50 DBS-paired blood samples assayed on the AU680 correlated with the CRP concentrations of capillary blood DBS and EDTA venous blood DBS (R^2^ = 0.99 and 0.99, respectively). The CYC within-assay imprecision (CV) was 3.7% and between-assay imprecision was 7.0%. The CYC concentrations of 30 DBS-paired serum and plasma samples assayed by ELISA correlated with the CYC concentrations of capillary blood DBS and EDTA venous blood DBS (R^2^ = 0.82 and 0.75, respectively).

### Supplement S3: Description of the SHARE multiplex performed at the Danish Center for Neonatal Screening, Department for Congenital Disorders, Statens Serum Institut, Copenhagen, Denmark

#### Samples analysis

Two 3.2mm disks from dried blood spot (DBS) samples were punched into each well of Nunc 96-well polystyrene microwell plates (#277143, Thermo Fisher Scientific) and stored wrapped in parafilm at −20°C until analyses. On the day of analyses, 130µL extraction buffer (PBS containing complete protease inhibitor cocktail, #11836145001; Roche Diagnostics) was added to each well, and the samples were incubated for 1 hr. at room temperature (RT) on a microwell shaker set at 900rpm. Extracts were analysed with a multiplex immunoassay using customized preprinted Meso-Scale plates (Meso-Scale Diagnostics (MSD), Rockville, MD, USA) coated with antibodies specific for ApoE4, BDNF, Clusterin (APOJ), EGF, IL-8, IL-12, IL-16, IL-18, MCP-1, and VEGF-A.

The ApoE4 assay was based on an ApoE4-specific antibody for capture (#M067-3, MBL International, Woburn, MA, USA) and an ApoE-pan-specific antibody (MSD) for detection using recombinant Human ApoE4 (#JM-4699, MBL International) as calibrator. The preprinted MSD plates were incubated at RT with blocker A (#R93BA) for 30 min, washed, and extracts were mixed 7:1 with 6x custom diluent (based on diluent 43, #R50AG, MSD) on the MSD plate. In addition, to artificially lower the signal of Clusterin, we added unlabeled Clusterin capture antibody (MSD) at a final concentration of 1ug/mL. Calibrators were diluted in diluent 7 (#R54BB), detection antibodies were diluted in diluent 3 (#R50AP, MSD).

Controls were made in-house from part of the calibrator solution in one batch, aliquoted in portions for each plate, and stored at −20°C until use. Extracts were incubated on the MSD plate for 2 hrs. at RT before washing and adding detection antibody following additional 2 hrs. incubation at RT. The samples were read on the QuickPlex SQ 120 (MSD) 4 min after adding 2x Read Buffer T (#R92TC, MSD). Analyte concentrations were calculated from the calibrator curves on each plate using 4PL logistic regression using the MSD Workbench software.

#### Analytical characterisation

Intra-assay variations were based on 24 replicate measurements of an internal control. Inter-assay variations were calculated from controls analysed in duplicate on each plate during the sample analysis, 212 plates in total. The lower limit of detections was calculated as 2.5 standard deviations above the average of repeated measurements of the zero calibrator. The higher detection limit was defined as the highest calibrator concentration. Detection limits and assay variation are outlined in Table S3.1:

**Supplementary Table S3.1:** Detection limits and assay variation biomarker set 2.

|  | Detection limit (pg/mL) |  | Coefficient of variation (CV%) |  |
| --- | --- | --- | --- | --- |
|  | Lower | Upper | Intra-assay variation | Inter-assay variation |
| ApoE4 | 3672 | 6,250,000 | 6.96 | 17.9 |
| BDNF | 12.19 | 12,500 | 14.6 | 10.7 |
| Clusterin (APOJ) | 4286 | 16,000,000 | 3.81 | 15.4 |
| EGF | 0.098 | 125 | 5.64 | 10.0 |
| IL-16 | 30.97 | 2880 | 7.38 | 12.5 |
| IL-18 | 1.072 | 2500 | 3.96 | 8.79 |
| IL-8 | 0.309 | 577 | 3.56 | 7.43 |
| IL-12 | 0.477 | 865 | 4.31 | 8.91 |
| MCP-1 | 0.573 | 577 | 5.05 | 7.05 |
| VEGF-A | 0.353 | 1210 | 3.95 | 7.41 |

**Abbreviations** ApoE4: Apolipoprotein E4; BDNF: Brain-derived neurotrophic factor; EGF: Epidermal growth factor; IL-16, IL-18; IL-8, IL-12: Interleukins -16, -18, -8, -12; MCP-1: Monocyte chemoattractant protein-1; VEGF-A: Vascular epithelial growth factor A

### Supplement S4: Description of the SHARE S-Plex analyses performed at the Danish Center for Neonatal Screening, Department for Congenital Disorders, Statens Serum Institut, Copenhagen, Denmark

#### Samples analysis

Two 3.2mm disks from dried blood spot (DBS) samples were punched into each well of Nunc 96-well polystyrene microwell plates (#277143, Thermo Fisher Scientific) and stored wrapped in parafilm at −20°C until analyses. On the day of analyses 90µL extraction buffer (PBS containing complete protease (#11836145001) and PhosSTOP phosphatase (#04906845001) inhibitor cocktails; Roche Diagnostics) was added to each well, and the samples were incubated for 1 hr. at room temperature on a microwell shaker set at 600rpm.

Extracts were analysed on two separate plates for pTau217 (#K151APFS) and Tau(total), GFAP and NfL, (#K15639S) on two consecutive days using the highly sensitive S-Plex platform following the suppliers’ instructions (Meso-Scale Diagnostics (MSD), Rockville, MD, USA). 25µL undiluted DBSS extract was added to the MSD plates containing 25µL blocking solution and left shaking at 700rpm. Calibrator curves and internal controls (made in-house from part of the calibrator solution in one batch) were prepared on Nunc 96-well polystyrene microwell plates and frozen prior to analysis. All pipetting was performed using a Biomek i5 robot (Beckman-Coulter, Ramcon, Denmark). The samples were read on the QuickPlex SQ 120 (MSD) immediately after adding the read buffer. Analyte concentrations were calculated from the calibrator curves on each plate using 4PL logistic regression using the MSD Workbench software.

#### Analytical characterisation

Intra-assay variations were based on 37 replicate measurements of an internal control on the same plate. Inter-assay variations were calculated from controls analysed in duplicate on each plate during the sample analysis, 45 plates in total. The lower limit (LLOD) is the average of the lower limits of detection of each plate defined as 2.5 standard deviations above the background signal. The higher detection limit (ULOD) was defined as the highest calibrator concentration. Detection limits and assay variation are shown in Table S4.1 below (and were also provided together with the data):

**Supplementary Table S4.1:** Detection limit and assay variation biomarker set 3.

| Analyte | Coefficient of variation (CV%) |  | Detection limit (fg/mL) |  |
| --- | --- | --- | --- | --- |
|  | Intra-assay | Inter-assay | Upper | Lower |
| **pTau217** | 22.8 | 19.3 | 500000 | 93.86 |
| **Tau(total)** | 7.91 | 8.73 | 100000 | 24.23 |
| **GFAP** | 15.0 | 13.4 | 557500 | 84.82 |
| **NfL** | 18.7 | 14.3 | 2831250 | 147.4 |

**Abbreviations** pTau217: phosphorylated tau at threonine 217, GFAP: Glial fibrillary acidic protein; NFL: Neurofilament light chain

### Supplement S5: Correction for field conditions and blood-specimen type

Field conditions have impacted all markers and vary considerably (Table S5.1). In biomarker set 1, storage time, spot size, and fieldwork conditions impact CHO, CYC and HDL in particular, and less so the other biomarkers. Blood-specimen type narrows the distribution since these influences are adjusted for, yet TRG remains almost unchanged (Figure S5.1). The means of CHO and HDL are shifted to lower biomarker values, while the reverse holds for CRP and THB. CYC is robust to fieldwork conditions and sample quality. HbA1c had to be corrected for change of the analysis instrument while the other conversion steps have negligible effects on the HbA1c biomarker value since HbA1c is a ratio measure.

For the biomarkers of set 2, we observed that shipment time significantly impacts the levels of all biomarkers; we consider the field conditions as covariates. Drying time has a significant effect on the levels of BDNF, Clusterin, EGF, and IL-16. Outside temperature significantly affects the levels of IL-16 and MCP-1 and only Clusterin is affected by humidity protection (closed bag). Finally, spot size has a significant effect on all biomarkers except IL-18. In our prior work^21,22^ we showed that temperature and blood spot size have a significant impact on ApoE4 protein measures. Therefore, these variables were considered as covariates in the models that analysed predictors of ApoE4 levels.

For the neurodegenerative biomarkers (set 3) we find that pTau217 and NfL are robust to survey conditions, while GFAP was sensitive to ambient temperature, drying time and whether the shipping bag was open or closed. Therefore, we correct for these survey conditions in the statistical analysis (Börsch-Supan et al.^23^). Huber et al. and Kalanko et al.^14,17^ report similar field impact on these markers.

**Supplementary Table S5.1:**
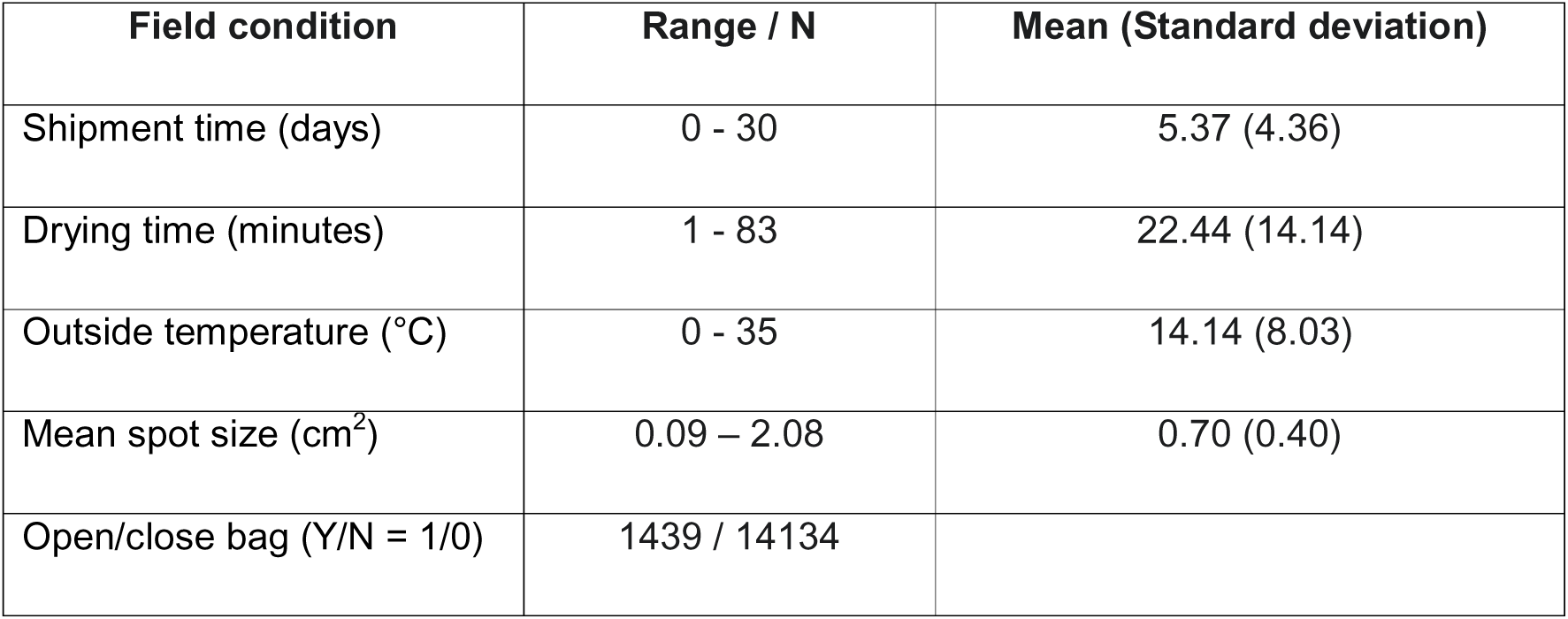
Fieldwork conditions.

| Field condition | Range / N | Mean (Standard deviation) |
| --- | --- | --- |
| Shipment time (days) | 0 - 30 | 5.37 (4.36) |
| Drying time (minutes) | 1 - 83 | 22.44 (14.14) |
| Outside temperature (°C) | 0 - 35 | 14.14 (8.03) |
| Mean spot size (cm<sup>2</sup>) | 0.09 – 2.08 | 0.70 (0.40) |
| Open/close bag (Y/N = 1/0) | 1439 / 14134 |  |

**Supplementary Figure S5.1:**
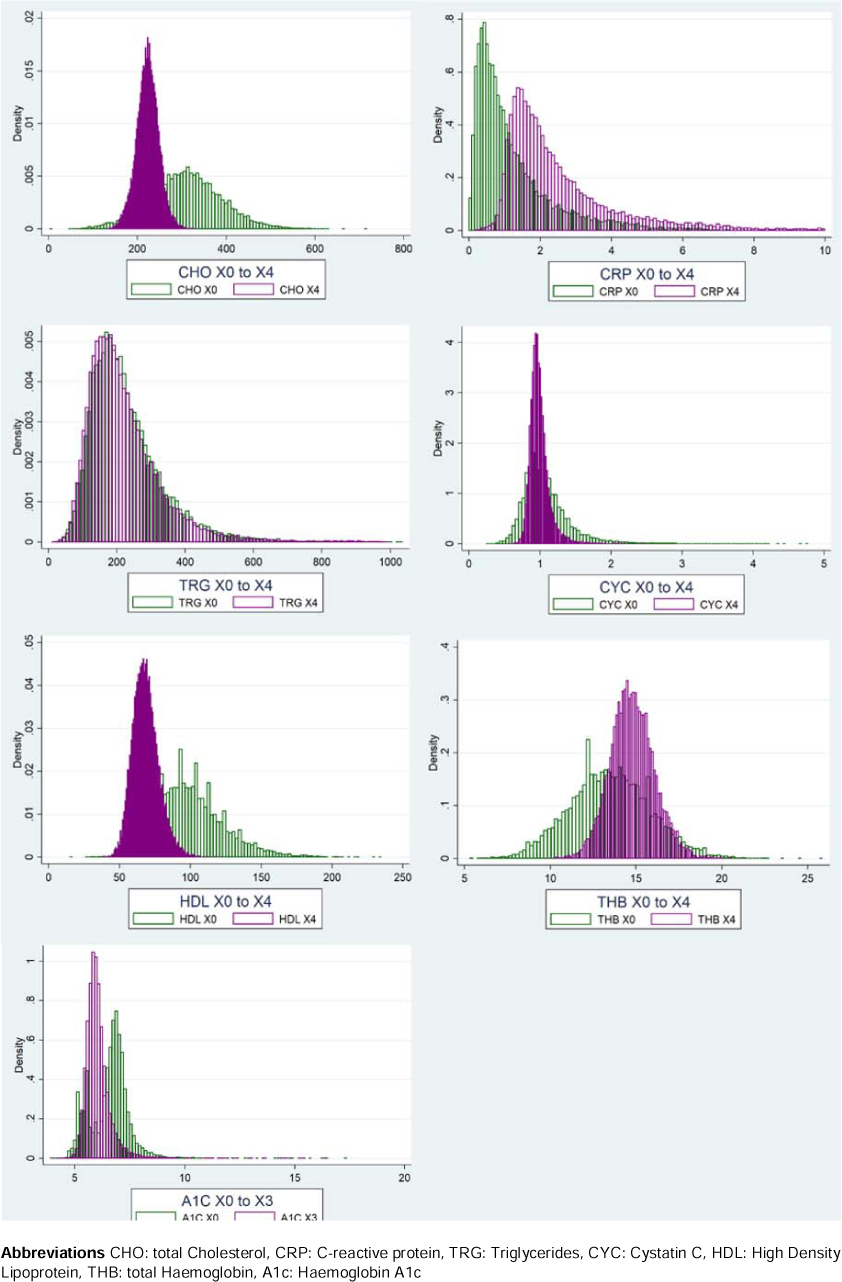
Distribution of raw and final biomarker data for marker set 1.

### Supplement S6: Descriptive statistics

**Supplementary Table S6.1:** Descriptive statistics by age, sex, education, BMI, and country.

|  | HbA1c | THb | HDL | CHO | TRG | CRP | CYC | ApoE4 | Clusterin | BDNF |
| --- | --- | --- | --- | --- | --- | --- | --- | --- | --- | --- |
|  | % | g/L | mg/dL | mg/dL | mg/dL | mg/L | mg/L | ng/mL | ng/mL | pg/mL |
| male | 6.07 | 15.09 | 65.59 | 219.5 | 206.1 | 2.53 | 0.977 | 58.03 | 1723.47 | 36.42 |
| female | 6.06 | 14.36 | 67.79 | 224.6 | 202.5 | 2.55 | 0.982 | 55.61 | 1778.42 | 41.27 |
| age<60 | 6.07 | 14.57 | 67.20 | 222.3 | 203.7 | 2.89 | 1.064 | 62.21 | 1785.81 | 36.64 |
| age 60-74 | 5.97 | 14.96 | 68.91 | 227.6 | 208.9 | 2.42 | 0.953 | 60.44 | 1791.61 | 40.28 |
| age>=75 | 6.08 | 14.65 | 66.39 | 221.5 | 203.5 | 2.55 | 0.982 | 55.09 | 1743.28 | 38.90 |
| educ1 | 6.07 | 14.68 | 66.25 | 222.1 | 206.8 | 2.88 | 1.012 | 52.80 | 1811.72 | 39.27 |
| educ2 | 6.03 | 14.87 | 68.82 | 228.4 | 209.1 | 2.51 | 0.976 | 65.44 | 1808.72 | 40.50 |
| educ3 | 6.07 | 14.61 | 66.11 | 220.7 | 202.1 | 2.46 | 0.972 | 49.21 | 1667.40 | 37.93 |
| bmi<24 | 5.88 | 14.61 | 70.77 | 227.4 | 186.9 | 2.15 | 0.970 | 59.80 | 1814.92 | 39.71 |
| bmi 24-29 | 6.02 | 14.92 | 67.71 | 227.1 | 210.8 | 2.52 | 0.984 | 57.12 | 1806.28 | 39.57 |
| bmi 30+ | 6.13 | 14.57 | 64.68 | 219.3 | 205.5 | 2.63 | 0.980 | 50.76 | 1652.20 | 38.19 |
| Sweden | 6.03 | 15.04 | 73.25 | 233.3 | 207.0 | 2.49 | 1.025 | 65.46 | 1942.82 | 40.39 |
| Estonia | 6.02 | 14.63 | 67.78 | 223.2 | 198.1 | 2.52 | 0.986 | 60.44 | 1672.40 | 43.18 |
| Denmark | 5.89 | 15.00 | 71.88 | 226.9 | 190.1 | 2.47 | 0.995 | 73.81 | 1767.44 | 45.16 |
| Germany | 6.07 | 14.76 | 67.49 | 224.3 | 204.4 | 2.53 | 0.988 | 64.60 | 1743.30 | 39.41 |
| Belgium | 5.97 | 15.00 | 69.60 | 222.5 | 201.6 | 2.45 | 0.992 | 61.15 | 1836.95 | 41.90 |
| France | 5.97 | 14.98 | 69.86 | 227.6 | 215.2 | 2.48 | 0.975 | 57.36 | 1786.45 | 39.78 |
| Switzerland | 5.98 | 15.00 | 72.20 | 231.4 | 213.0 | 2.40 | 1.012 | 70.03 | 1977.12 | 39.81 |
| Slovenia | 6.15 | 14.97 | 67.03 | 222.3 | 208.7 | 2.61 | 0.985 | 58.70 | 1814.73 | 39.50 |
| Italy | 6.09 | 14.55 | 63.96 | 217.8 | 198.3 | 2.65 | 0.991 | 37.83 | 1751.02 | 39.11 |
| Spain | 6.18 | 14.22 | 61.60 | 214.7 | 196.7 | 2.56 | 0.940 | 52.53 | 1634.37 | 34.58 |
| Greece | 6.32 | 13.91 | 61.58 | 218.7 | 203.6 | 2.45 | 0.960 | 46.92 | 1703.69 | 37.90 |
| Israel | 6.13 | 14.16 | 60.92 | 203.1 | 199.4 | 2.57 | 0.939 | 50.09 | 1499.91 | 37.35 |

**Supplementary Table S6.1 (continued): Descriptive statistics by age, sex, education, BMI, and country**
|  | EGF | IL-16 | IL-8 | IL-18 | IL-12 | MCP-1 | VEGF-A | Tau total | GFAP | NfL | pTau217 |
| --- | --- | --- | --- | --- | --- | --- | --- | --- | --- | --- | --- |
|  | pg/mL | pg/mL | pg/mL | pg/mL | pg/mL | pg/mL | pg/mL | pg/mL | pg/mL | pg/mL | pg/mL |
| male | 2.94 | 326.7 | 2.88 | 119.3 | 11.04 | 46.71 | 13.81 | 11.27 | 1.19 | 3.34 | 2.14 |
| female | 3.29 | 324.6 | 5.05 | 100.0 | 12.63 | 43.02 | 14.38 | 10.34 | 1.33 | 2.78 | 2.09 |
| age<60 | 2.94 | 337.1 | 3.21 | 125.6 | 12.37 | 44.14 | 14.53 | 10.25 | 1.90 | 4.28 | 2.29 |
| age 60-74 | 3.19 | 332.8 | 2.71 | 103.1 | 11.15 | 45.26 | 13.80 | 10.72 | 1.04 | 2.40 | 2.06 |
| age>=75 | 3.13 | 323.3 | 4.46 | 109.5 | 12.11 | 44.54 | 14.19 | 10.77 | 1.32 | 3.17 | 2.11 |
| educ1 | 3.08 | 325.6 | 3.18 | 108.5 | 12.54 | 44.15 | 14.44 | 10.74 | 1.50 | 3.13 | 2.20 |
| educ2 | 3.16 | 334.9 | 2.92 | 108.1 | 12.21 | 45.17 | 14.13 | 11.01 | 1.20 | 3.01 | 2.14 |
| educ3 | 3.16 | 318.2 | 5.67 | 109.2 | 11.23 | 44.68 | 13.88 | 10.31 | 1.27 | 2.99 | 2.01 |
| bmi<24 | 3.16 | 330.7 | 3.09 | 102.2 | 11.88 | 44.24 | 13.63 | 9.80 | 1.39 | 3.00 | 2.01 |
| bmi 24-29 | 3.09 | 326.6 | 2.80 | 109.1 | 12.09 | 44.49 | 14.09 | 10.72 | 1.19 | 2.82 | 2.10 |
| bmi 30+ | 3.18 | 320.9 | 6.25 | 112.3 | 11.77 | 45.16 | 14.49 | 12.00 | 1.32 | 3.60 | 2.34 |
| Sweden | 2.94 | 336.5 | 2.70 | 113.8 | 12.61 | 46.62 | 13.62 | 11.10 | 1.38 | 2.81 | 1.99 |
| Estonia | 3.46 | 319.7 | 3.15 | 109.8 | 12.00 | 46.96 | 14.02 |  |  |  |  |
| Denmark | 3.60 | 341.7 | 2.93 | 108.2 | 12.59 | 47.58 | 14.31 | 10.85 | 1.22 | 2.85 | 2.26 |
| Germany | 3.15 | 328.7 | 2.95 | 108.2 | 11.66 | 45.59 | 14.19 | 10.68 | 1.26 | 3.10 | 2.10 |
| Belgium | 3.22 | 342.8 | 2.79 | 112.7 | 11.91 | 47.94 | 14.16 |  |  |  |  |
| France | 3.33 | 331.9 | 2.90 | 107.1 | 12.00 | 43.54 | 14.47 |  |  |  |  |
| Switzerland | 3.08 | 340.1 | 2.67 | 109.9 | 12.33 | 47.44 | 14.16 |  |  |  |  |
| Slovenia | 2.90 | 329.6 | 2.68 | 109.5 | 12.47 | 46.60 | 14.16 |  |  |  |  |
| Italy | 3.24 | 316.7 | 3.28 | 107.7 | 12.41 | 43.67 | 13.77 |  |  |  |  |
| Spain | 2.59 | 317.1 | 12.25 | 104.3 | 11.71 | 43.68 | 14.27 |  |  |  |  |
| Greece | 2.91 | 311.4 | 2.61 | 126.1 | 10.70 | 42.44 | 13.65 |  |  |  |  |
| Israel | 3.08 | 267.4 | 7.40 | 106.7 | 11.45 | 41.91 | 13.70 |  |  |  |  |

**Abbreviations** HbA1c: Haemoglobin A1c (glycosylated haemoglobin); THb: total Haemoglobin; HDL: High density lipoprotein; CHO: total Cholesterol; CYC: Cystatin C; ApoE4: Apolipoprotein E4; BDNF: Brain-derived neurotrophic factor; EGF: Epidermal growth factor; IL-16, IL-8; IL18, IL12: Interleukins 16, 8, 18, 12; MCP-1: Monocyte chemoattractant protein-1; VEGF-A: Vascular epithelial growth factor-A; GFAP: Glial fibrillary acidic protein; NFL: Neurofilament light chain; pTau217: phosphorylated tau at threonine 217
Age in years; Education: Participants were grouped into three educational categories according to the International Standard Classification of Education (ISCED). educ1: low education (ISCED 0–2, pre-primary through lower secondary), educ2: medium education (ISCED 3–4, upper secondary and post-secondary non-tertiary) and educ3: high education (ISCED 5–6, tertiary degrees). https://ec.europa.eu/eurostat/statistics-explained/index.php?title=International_Standard_Classification_of_Education_(ISCED); BMI in kg/m<sup>2</sup>;

